# Acceptability and implementation of digital mental health supports for marginalised young people across Ireland: A mixed-methods study

**DOI:** 10.64898/2026.08.08.26359861

**Authors:** Carmen Kealy, Adele Mc Loughlin, Alba Madrid-Cagigal, Siobhan O’Neill, Gary Donohoe, Maurice D. Mulvenna, Margaret M. Barry

## Abstract

Digital mental health tools are increasingly promoted as scalable supports for young people, yet implementation remains inconsistent, particularly for marginalised youth. Acceptability and usability are key determinants of successful adoption, but little is known about how these factors shape engagement across diverse youth populations.

The aim of the study was to examine the acceptability, usability, and implementation potential of 11 evidence-based digital mental health tools among marginalised young people across the Republic of Ireland (ROI) and Northern Ireland (NI).

A mixed-methods design integrated baseline surveys (n = 38), a two-week trial of digital tools delivered through a co-designed Google Site, online workshops/individual interviews (n = 22), and a final usability and engagement survey (n = 24). Usability was assessed using the System Usability Scale (SUS), engagement using the Twente Engagement with E-Health Technologies Scale (TWEETS), and mental wellbeing using the Short Warwick–Edinburgh Mental Well-Being Scale (SWEMWBS). Qualitative data were analysed thematically and mapped to the Consolidated Framework for Implementation Research (CFIR).

Only two tools exceeded the SUS usability benchmark. Engagement was moderate overall, with one tool achieving the highest engagement despite lower usability. SWEMWBS scores indicated moderate baseline mental wellbeing. Thematic analysis identified five acceptability themes: credibility and trust; accessibility and ease of use; positive content supporting emotional regulation; personalisation and self-monitoring; and engagement and habit formation. CFIR analysis highlighted usability, institutional trust, cultural relevance, and emotional needs as core implementation determinants. Digital literacy was high and supported engagement, and usability remained a critical gateway to implementation. Designers and commissioners of digital mental health tools should ensure that supports are simple, trustworthy, culturally relevant, and youth-centred to enable adoption among marginalised young people. Implementation strategies are needed that will co-design with diverse youth communities and prioritise youth work settings as well as governance clarity.

**Author Summary:** Digital mental health tools are often promoted as a way to support young people who face barriers to traditional services, yet we still know little about how acceptable or usable these tools are for diverse youth communities. In this study, we worked with marginalised young people aged 18–25 across the Republic of Ireland and Northern Ireland to understand how they experience a range of evidence-based digital mental health supports. Using surveys, workshops, interviews, and a two-week trial of eleven tools, we explored what makes digital supports feel trustworthy, easy to use, engaging, and relevant.

We found that usability is a critical gateway: young people are far more likely to use tools that are simple, intuitive, and free of unnecessary steps. Trust in recognised health or youth organisations also strongly shaped engagement. Many participants used digital tools for emotional regulation—especially those offering positive or calming content—and valued features that helped them track their mood or personalise their experience. However, complex interfaces, paywalls, and unclear credibility reduced uptake.

Our findings show that digital mental health supports must be youth-centred, culturally relevant, and easy to navigate to be successfully implemented, particularly for marginalised young people.

## Introduction

Digital mental health supports are increasingly positioned as scalable, low-threshold interventions capable of expanding access to mental health care for young people [1.2]. As the field evolves beyond traditional app-based interventions to encompass AI-enabled tools, virtual reality supports, and blended digital–human models, questions of implementation, safety, and sustained engagement have become more prominent [2]. Across the Republic of Ireland (ROI) and Northern Ireland (NI), policy frameworks such as *Sharing the Vision* [3], the *NI Mental Health Strategy 2021–2031* [4], and *Digital for Care* [5] emphasise the potential of digital tools to complement overstretched services, reduce waiting times, and provide flexible, youth-centred support. This aligns with broader international policy, including the WHO *Global Strategy on Digital Health* and youth eHealth frameworks, which advocate for integrating digital technology into national health portfolios to bridge the mental health treatment gap among young users [6]. Recent evidence from global health systems further highlights how digital transformation can reduce structural access barriers when supported by governance, infrastructure, and equity-focused implementation [7].

Yet despite this policy momentum, implementation remains inconsistent, and engagement with digital mental health tools is highly variable [8]. Young people, particularly those who are marginalised, continue to report barriers related to usability, cultural relevance, trust, and digital literacy [9]. Systematic evidence shows that these barriers are not incidental but structural and predictable, encompassing issues such as low perceived relevance, privacy concerns, lack of personalisation, and insufficient human support, all of which shape uptake and sustained use [8]. Ireland’s largest national youth mental health study, the *My World Survey 2* [1], underscored the urgency behind these digital initiatives, documenting a nationwide escalation in anxiety and depression alongside declines in protective psychological assets such as self-esteem, optimism, and resilience.

Marginalised youth — including migrants, Travellers, LGBTQ+ young people, rural youth, young people not in education, employment or training (NEET), and those with disabilities or neurodivergence — face disproportionate structural and social barriers to accessing mental health support [11]. Digital tools are often assumed to mitigate these inequities by offering anonymity, immediacy, and autonomy [7]. However, emerging evidence suggests that digital mental health interventions frequently fail to meet the needs of diverse youth populations, with limited representation, complex interfaces, and insufficient cultural adaptation [12–14]. Even advanced technologies such as AI-driven chatbots and VR-based interventions face persistent challenges related to algorithmic transparency, data governance, and clinical validation, which can further erode trust among marginalised groups [2], Engagement is also shaped by young people’s perceptions of credibility, institutional trust, and the emotional resonance of digital content, which act as vital bridges in digital environments and influence user choice and sustained long-term attachment [15].

Acceptability and usability are central determinants of implementation success [2, 8]. Consistent with previous digital mental health research, usability is defined as the degree to which digital mental health supports are perceived as easy, efficient, and satisfactory to use, including aspects of engagement, technical functionality, and ease of interaction [1, 14]. Acceptability is defined as the extent to which these supports are viewed as satisfactory, appropriate, and valuable by users, as reflected in satisfaction, perceived suitability, and intentions to adopt and continue using the intervention [16,17]. The System Usability Scale (SUS) [18] and the Twente Engagement with E-Health Technologies Scale (TWEETS) [19] provide robust measures of how young people experience digital tools, while implementation frameworks such as the Consolidated Framework for Implementation Research (CFIR) [20] help explain how intervention characteristics, individual needs, and contextual factors shape adoption [21]. Yet few studies integrate these quantitative and qualitative dimensions, and none have examined acceptability and implementation across an all-island, marginalised youth sample.

This study addresses this gap by evaluating the usability, acceptability, and implementation potential of 11 evidence-based digital mental health tools among marginalised young people aged 18–25 across ROI and NI. Using a mixed-methods design, we combined SUS and TWEETS scores with qualitative thematic analysis and CFIR-aligned interpretation to generate a comprehensive understanding of what makes digital tools acceptable, usable, and implementable for diverse youth populations. This study offers one of the first all-island analyses of digital mental health implementation for marginalised youth in Ireland and provides actionable insights for policymakers, service providers, and developers seeking to build trustworthy, youth-centred digital ecosystems.

## Materials and methods

### Study design

A mixed-methods sequential design integrated quantitative usability and engagement measures with qualitative thematic analysis on acceptability and CFIR-aligned implementation interpretation. The study comprised four phases: a baseline survey capturing demographics and mental wellbeing using the Short Warwick-Edinburgh Mental Wellbeing Scale (SWEMWBS)[22]; a two-week trial of digital mental health tools delivered through a co-designed Google Site; online workshops and targeted interviews exploring usability, trust, cultural relevance, emotional impact, and implementation preferences; and a final survey assessing usability, engagement, acceptability, trust, and implementation preferences. This sequential approach enabled triangulation across behavioural, experiential, and implementation-focused data sources, supporting a comprehensive assessment of acceptability and implementation potential.

### Participants and recruitment

The recruitment for this study took place between August 2025 and January 2026. Participants were young people aged 18–25 residing in the Republic of Ireland (ROI) or Northern Ireland (NI). Recruitment prioritised marginalised youth, including migrants, asylum seekers, Traveller youth, LGBTQ+ young people, rural youth, NEET young people, and those with disabilities or neurodivergence. When the study flyer was circulated initially on social media, recruitment generated several responses from young people not resident in Ireland or otherwise not meeting the selection criteria for this study; these were removed during data cleaning, after which recruitment proceeded exclusively through verified youth organisations and trusted gatekeepers to ensure data integrity and participant safety.

A total of 38 participants completed the baseline survey, 22 took part in online workshops or interviews, and 24 completed the final survey. Participants represented diverse ethnic, cultural, and social backgrounds, consistent with the project’s focus on equity, inclusion, and culturally grounded participation

#### Ethics statement

This study received ethical approval from an accredited university research ethics committee. Prior to taking part in surveys, workshops, interviews, and the digital tool trial, all participants provided informed consent electronically via Jisc Online Surveys (–a secure, GDPR-compliant platform widely utilized across UK higher education). Participation was voluntary, and all data were anonymised in accordance with GDPR and institutional ethical guidelines. Additional safeguards were implemented to protect marginalised youth, including verified recruitment through trusted organisations and removal of fraudulent submissions to ensure participant safety and data integrity.

#### Digital tools evaluated

Eleven evidence-based digital mental health tools were selected through a co-design process with the project’s Youth Reference Group. Candidate tools were identified from an initial pool of 94 digital mental health resources curated through the ORCHA app library [23], other online sources, and suggestions from Youth Reference Group members. The research team checked the evidence base for all selected resources, including relevance to young people, accessibility, cost, and suitability for use in Ireland. Youth Reference Group members then reviewed and prioritised tools based on perceived usefulness, usability, relevance to everyday mental health needs, and likelihood of engagement. The final suite spanned information and signposting, mood tracking and emotional regulation, positive content, and skills-based support (Table 1). Participants accessed all tools through a co-designed Google Site, ensuring consistent navigation and standardised access. Usage was self-directed to reflect real-world engagement.

**Table 1.**
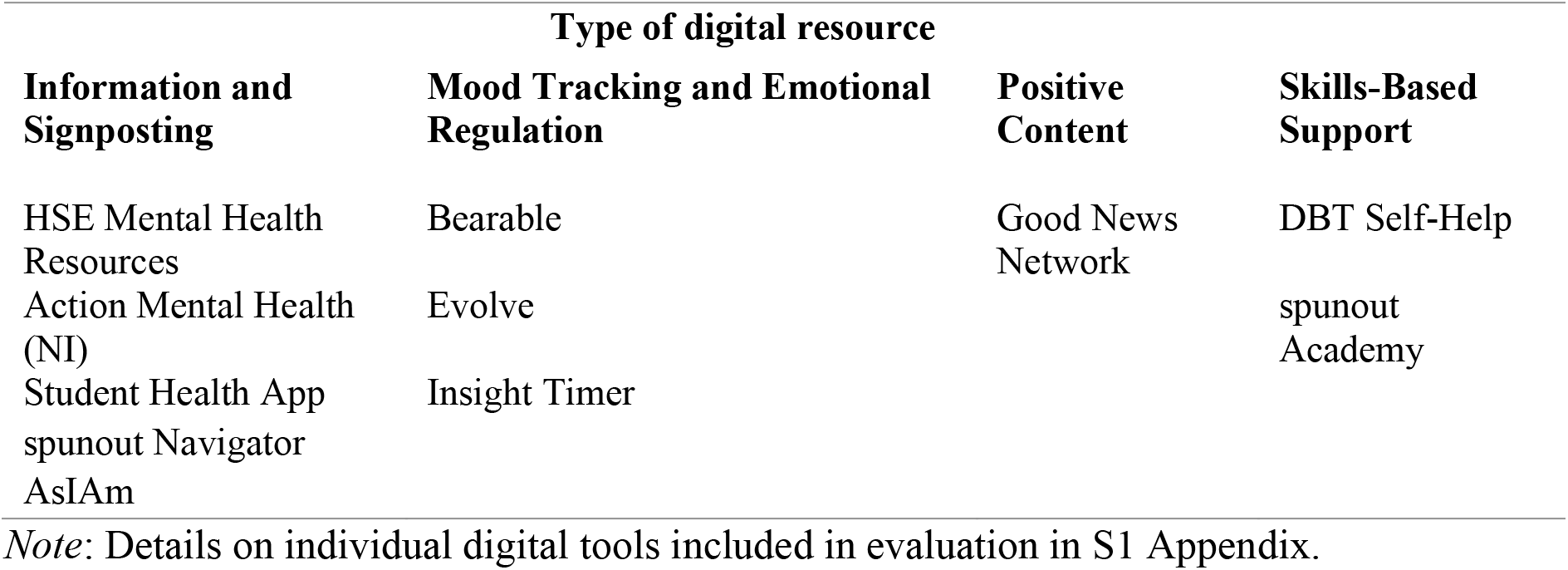
List of digital tools included in evaluation.

### Data collection

#### Baseline survey

The baseline survey captured demographic characteristics and mental well-being using the Short Warwick-Edinburgh Mental Wellbeing Scale (SWEMWBS).

#### Two-week tool trial

Participants accessed a dedicated Google Site hosting the selected eleven tools and were encouraged via email reminders to use them over a two-week period. Usage was self-directed to reflect real-world engagement patterns.

#### Workshops/individual interviews

Following completion of the baseline survey, participants attended a facilitated workshop or an interview, exploring their existing experiences with digital mental health supports and their expectations regarding the study tools. Guided discussion focused on perceptions of trustworthiness, usability, accessibility, cultural relevance, and factors likely to influence engagement. Workshops and interviews were audio-recorded and transcribed verbatim for analysis. Participants selected pseudonyms to support anonymity during discussion. Additional detail on the question prompts is provided in S2 Appendix.

#### Final survey

The final survey assessed mental wellbeing using the Short Warwick–Edinburgh Mental Wellbeing Scale (SWEMWBS), usability through the System Usability Scale (SUS), and engagement using the Twente Engagement with E-Health Technologies Scale (TWEETS). It also captured acceptability, trust, and implementation preferences through open-ended questions.

#### Measures

#### SWEMWBS (Baseline and Final survey)

The Short Warwick–Edinburgh Mental Wellbeing Scale (SWEMWBS) [22] was used to assess participants’ mental wellbeing. The scale consists of 7 items capturing aspects of positive mental functioning, including optimism, clear thinking, and feeling close to others. Participants rated each item on a 5-point Likert scale ranging from 1 (none of the time) to 5 (all of the time), with higher total scores indicating greater mental wellbeing. Scores are typically transformed to metric scores, using a validated conversion table to allow comparison with population norms.

#### System Usability Scale (SUS) (Final survey)

Perceived usability was assessed using the System Usability Scale (SUS) [18], a widely used and validated 10-item measure of system usability. Items are scored on a five-point Likert scale and converted to a total score ranging from 0–100, with higher scores representing higher usability. A score of 68 is commonly considered the industry benchmark for acceptable usability.

#### TWEETS Engagement Scale (Final survey)

Engagement with digital tools was assessed using the TWente Engagement with E-Health Technologies Scale (TWEETS) [19], a 9-item measure assessing behavioural, cognitive, and affective engagement with digital health tools. Responses are recorded on a five-point Likert scale, with higher scores indicating greater engagement. For this study, the items were adapted by replacing “technology” with “app/website”.

#### Acceptability and implementation questions

Acceptability and implementation preferences were assessed through brief questions on information sought about the source and functioning of digital mental health tools, the importance of data protection, perceived advantages over other supports, reasons for choosing alternative options, features that would encourage continued use, sustained use after the study, perceived interest among peers, and views on youth involvement in co-design, with full item wording provided in S3 Appendix.

### Data analysis

#### Quantitative analysis

Quantitative analyses were conducted using SPSS version 31 [24]. Descriptive statistics (means, standard deviations, frequencies and percentages) were calculated for participant characteristics and survey measures. Mean SUS and TWEETS scores were computed to assess perceived usability and engagement. Engagement patterns were compared across tools, and mean differences were interpreted in relation to usability and qualitative findings. Quantitative findings were subsequently integrated with qualitative findings to provide a comprehensive understanding of acceptability and implementation.

#### Qualitative analysis

Audio-recorded workshop discussions and individual interviews were transcribed verbatim and analysed alongside open-ended survey responses using reflexive thematic analysis following the approach of Braun and Clarke [25]. Analysis was primarily inductive but informed by the Consolidated Framework for Implementation Research (CFIR), allowing findings to be interpreted in relation to implementation determinants. Coding was iterative and interpretive, with themes refined through team discussion to ensure coherence with CFIR constructs across intervention characteristics, individual needs, and contextual influences. Final themes captured participants’ experiences relating to credibility and trust, accessibility and ease of use, positive content as emotional regulation, personalisation and self-monitoring, and engagement and habit formation.

#### CFIR mapping

Themes were mapped onto four CFIR domains—Intervention Characteristics, Outer Setting, Characteristics of Individuals, and Process—which enabled interpretation of acceptability and engagement within an implementation science framework.

The research team brought diverse disciplinary backgrounds in psychology, health promotion, computing, and youth mental health. Co-design with the Youth Reference Group informed tool selection and study design, supporting relevance and cultural grounding. Reflexive discussions throughout analysis helped identify potential biases, particularly regarding assumptions about digital literacy, cultural relevance, and institutional trust. These reflexive practices strengthened the credibility and transparency of the findings.

## Results

A descriptive overview of participant demographics and mental wellbeing is provided in Tables 2–3, followed by key metrics from the co-designed Google site (Table 4). The subsequent findings integrate quantitative and qualitative data within a unified thematic structure. Quantitative indicators—including SUS usability scores and TWEETS engagement ratings— are presented within the relevant themes, where they complement and contextualise qualitative insights. Given the limited quantitative sample size, qualitative data, including open-ended responses from the final survey, provide the primary explanatory depth. Integrated findings are organised through reflexive thematic analysis, capturing core dimensions of acceptability and implementation.

**Table 2.** Characteristics of Study Participants.

| Data Collection type | Total (n)<br>ROI & NI | Region & Gender (n) |  | Age Range | Ethnicity<br>White, British Isles % | Mental Health Condition % | Identifying as Migrant/ Immigrant % |
| --- | --- | --- | --- | --- | --- | --- | --- |
|  |  | ROI | NI |  |  |  |  |
| Baseline Survey | 38 | F=14;<br>M=1;<br>Other=1 | F=5;<br>M=16;<br>Other=1 | 18-25<br>(M = 21.34) | 50.00 | 34.21 | 18.42 |
| Final Survey | 24 | F=6;<br>M=0 | F=5;<br>M=13 | 18-25<br>(M = 21.42) | 33.33 | 20.81 | 25.00 |
| **Pilot** | 2 | F=1 | F=1 | 21-23<br>(M = 22.00) | 100.00 | 50.00 | 0 |
| **Workshop 1** | 11 | 0 | M=9 | 19-25<br>(M = 21.18) | 0 | 0 | 36.36 |
| **Workshop 2** | 7 | F=1 | F=2;<br>M=4 | 20-24<br>(M = 22.40) | 20.00 | 0 | 20.00 |
| **Individual Interviews** | 4 | F=4 | 0 | 20-23<br>(M = 21.25) | 100.00 | 25.00 | 0 |

*Note.* Participants resided in ROI (Republic of Ireland) or NI (Northern Ireland). F=Female; M=Male; Gender Other = Non-binary (n=1) and Gender Fluid (n=1). Other ethnicities included Black, Black British, Caribbean or African; Mixed or multiple ethnic groups and White (Gypsy or Irish Traveller; Roma). Migrants include refugees and asylum seekers. Those identifying as part of the LGBTQIA+ community were; Baseline (n=7), Final (n=1), Workshop 2 (n=1), Interview (n=1).

**Table 3.** Participant mental wellbeing across baseline and final surveys using the Short Warwick–Edinburgh Mental Well-Being Scale (SWEMWBS).

|  | **Baseline**<br>**NI & ROI** | **Baseline**<br>**NI** | **Baseline**<br>**ROI** | **Final**<br>**NI & ROI** | **Final**<br>**NI** | **Final**<br>**ROI** |
| --- | --- | --- | --- | --- | --- | --- |
| **Mean** | 20.05 | 18.32 | 22.44 | 25.83 | 25.83 | 25.83 |
| **Range** | 10-30 | 11-27 | 10-30 | 18-35 | 22-31 | 18-35 |
| **SD** | 5.23 | 4.54 | 5.32 | 3.61 | 2.18 | 6.62 |

*Note:* NI=Northern Ireland; ROI=Republic of Ireland

**Table 4.** Downloads/access of mental health supports by percentage.

| **Digital Health Resource** | **% Popularity** |
| --- | --- |
| Student Health App | 19 |
| Bearable | 14 |
| Good News Network | 12 |
| Evolve | 11 |
| DBT Self-Help | 8 |
| spunout Navigator | 8 |
| Action Mental Health | 7 |
| Insight Timer | 6 |
| AsIAm | 6 |
| spunout Academy | 5 |
| HSE Mental Health | 4 |
|  | 100 |

The study cohort comprised young people aged 18 to 25 years (Table 2), residing across the ROI and NI, purposefully sampled to explore the acceptability of digital mental health tools among diverse and marginalised youth. The baseline survey (n = 38) showed a somewhat even distribution of participants identifying as female (50%) and male (44.7%), alongside individuals identifying as non-binary or gender fluid (5.3%). A subset of these participants completed a final evaluative survey (n = 24; 45.8% female, 54.2% male) where females were evenly split across NI and ROI and all male participants resided in NI. However, disparities were evident between the ROI sample which comprised higher female participation and higher overall attrition, whereas the NI sample demonstrated lower dropout and greater male representation. Qualitative input through a pilot framework, online workshops and targeted interviews (n = 22), typified the study’s focus on underrepresented or vulnerable populations. A notable portion of the baseline sample (34.2%) and final survey (20.8%) reported living with an established mental health condition although this dropped to zero among workshop participants. Single recorded health complexities across the wider cohort included mobility issues, vision impairments, long term illness, ADHD and Autism spectrum disorder. Furthermore, LGBTQ+ identification stood at 18% in the baseline survey adjusting to 4% by the final survey. While the majority of Pilot (50%) and Interview participants (100%) identified as White, (Irish, English, Welsh, Scottish, Northern Irish or British), the Workshops captured a large representation of migrants (including refugees and asylum seekers) and immigrants, the majority of which identified as Black, Black British, Caribbean or African or of Mixed or multiple ethnic groups. Functionally, the cohort were highly active, with many balancing combined roles, across education, employment, and volunteering.

Participant mental wellbeing was assessed at baseline (n = 38) and in the final survey (n = 24) using the Short Warwick–Edinburgh Mental Well-Being Scale (SWEMWBS) (Table 3).

Participant mental wellbeing was evaluated across the baseline (n = 38) and final surveys (n = 24) using the SWEMWBS (Table 3). Overall, mean scores reflected stable, moderate levels of mental wellbeing throughout the study, moving from baseline (M = 20.05, SD = 5.23) to final evaluation (M = 25.83, SD = 3.61). While the quantitative sample size limits formal statistical inferencing, these scores establish important cohort baseline context, complementing the qualitative feedback on how digital tools supported light-touch emotional regulation and daily wellbeing maintenance.

The study also collected key metrics on the co-designed Google Site of digital mental health supports, including the most popular supports (ranked list by % popularity in Table 4).

Each visitor to the co-designed Google Site viewed approximately 8 of the 11 supports. 13% of the visitors downloaded one or more supports, with an average session time appraising and downloading them of 5 minutes 37 seconds.

### Thematic analysis of acceptability and engagement

Five interconnected themes captured the core dimensions of acceptability: credibility and trust; accessibility and ease of use; positive content as emotional regulation; personalisation and self-monitoring; and engagement and habit formation. These themes were not discrete categories but overlapping influences that shaped how young people navigated, interpreted, and integrated digital tools into their daily lives.

#### Credibility and trust

Credibility emerged as a foundational determinant of acceptability. Young people consistently evaluated tools through the lens of who created them, who endorsed them, and whether they aligned with recognised health or youth-support institutions. Tools associated with the HSE, Action Mental Health, NHS, or universities were prioritised for their “clinical validation” and perceived as inherently safer and more legitimate. Participants described these tools as, “credible and backed up by research,” “more reliable,” and “less likely to be misleading.” Alongside formal credibility, peer perspectives functioned as powerful informal validators. Youth-led design, testimonials “of others who have used the service”, and content created by, “people my age” increased relatability and trust. Participants emphasised that “positive ratings from people” made tools feel more authentic and less clinical. In contrast, commercial apps elicited cautious engagement. Concerns centred on profit motives, data harvesting, and manipulated statistics. However, credibility checks were not universal. For positive-content tools such as Good News Network, emotional benefit outweighed evidence considerations. Participants openly acknowledged that they “didn’t check credibility” because the content made them feel better, illustrating how emotional resonance can override critical appraisal.

#### Accessibility and ease of use

Usability was the most consistent determinant of acceptability across the dataset. Participants valued tools that were simple, intuitive, and required minimal cognitive effort. Tools described as “easy to navigate,” “not overwhelming,” and “accessible anywhere” were consistently preferred. These qualitative accounts aligned closely with SUS patterns (Table 5), where only two tools exceeded the usability benchmark of 68. Tools that opened immediately, required no sign-up, and used clear, concise language were perceived as more equitable and more aligned with young people’s needs.

**Table 5.** System Usability Scale (SUS) Mean Results, ranked evaluation of Digital Platforms.

| Rank | Resource | Mean SUS Score |
| --- | --- | --- |
| 1 | spunout Navigator | 71.88 |
| 2 | Student Health App | 69.58 |
| 3 | Action Mental Health | 65.00 |
| 4 | HSE Mental Health | 63.33 |
| 5 | Bearable | 62.81 |
| 6 | spunout Empathy Online (unit within spunout Academy) | 57.50 |

Barriers such as long pages, excessive options, repeated clicks, complex navigation or “school-like” interfaces reduced engagement. Participants described these tools as “too many clicks,” “overwhelming,” or “dragging on,” which discouraged continued use. Cost and paywalls were also significant barriers. Free tools were perceived as more accessible and more aligned with the realities of young people’s financial constraints. The pocket therapist lends 24/7 availability, thus removing the “scheduling barrier”, and provides immediate support during acute episodes, such as a “2 a.m. panic attack.” These findings emphasise that sustaining long-term youth engagement centres on removing both operational friction and financial barriers.

#### Positive content supporting emotional regulation

Across the sample, digital tools were used primarily as light-touch emotional coping supports rather than clinical interventions. Positive-content platforms such as Good News Network were valued for providing “instant positivity,” distraction, and relief from negative news cycles. Participants described these tools as mood-lifting, grounding, and emotionally protective, particularly during stressful periods.

This emotional function helps explain why some tools with lower usability still achieved high engagement. Bearable (M = 25.88) and Good News Network (M = 25.57) were used frequently because they offered immediate affective benefit. Young people prioritised emotional resonance over interface quality when a tool provided comfort, distraction, or a sense of relief. However, participants also recognised the limitations of these tools, noting that they were not substitutes for structured therapeutic support. This distinction underscores the importance of positioning digital tools as part of a broader ecosystem of supports rather than as standalone solutions.

#### Personalisation and self-monitoring

Personalisation enhanced relevance and supported self-insight. Participants valued tools that allowed them to track mood, tailor content, adjust complexity, or reflect on emotional patterns. Mood-tracking apps such as Bearable were described as helping users “see patterns” from their “daily check-ins” and understand triggers or fluctuations across the week. These features bridged the information gap for young adults, supported self-awareness and provided a sense of agency in managing emotional wellbeing.

Youth-specific or culturally relevant content, particularly Irish or UK examples, was perceived as more relatable and trustworthy. Participants emphasised that tools reflecting their identities, contexts, or cultural references felt more meaningful and less generic. However, personalisation had limits. Excessive options or complex tracking features created cognitive overload, particularly for neurodivergent users. Participants described feeling overwhelmed by tools that required too many decisions or too much input. This balance between flexibility and simplicity was central to perceived usefulness.

#### Engagement and habit formation

Sustained engagement was shaped by a combination of emotional payoff, simplicity, and behavioural reinforcement. Reminders, streaks, short tasks, in-app rewards and predictable interfaces supported routine use and helped embed tools into daily life. Participants described notifications as helpful prompts that encouraged consistent check-ins. Over 41% of participants reported that gamification and rewards incentivise continued use of tools. These behavioural rewards help establish daily habits, reducing the perceived effort of mental health maintenance and reframing mental health care as achievable “wins.” Variety and novelty also supported attention, particularly for participants with ADHD, who valued tools offering multiple content types or formats. Participant engagement with digital mental health resources was assessed using the TWente Engagement with Ehealth Technologies Scale (TWEETS) (Table 6).

**Table 6.** Participant engagement with Digital Health Resources in the previous 4 weeks using the TWente Engagement with Ehealth Technologies Scale (TWEETS).

| Digital Health Resource | Overall TWEET Scores |  |  |  | Behavioural Engagement |  |  | Cognitive Engagement |  |  | Affective Engagement |  |  |
| --- | --- | --- | --- | --- | --- | --- | --- | --- | --- | --- | --- | --- | --- |
|  | n | Range | Mean | SD | Range | Mean | SD | Range | Mean | SD | Range | Mean | SD |
| Spunout Navigator | 5 | 18 - 26 | 22.40 | 3.36 | 4-11 | 7.20 | 2.77 | 7-8 | 7.20 | 0.45 | 7-9 | 8.00 | 0.71 |
| Bearable | 5 | 18 - 26 | 23.40 | 3.21 | 7-10 | 8.60 | 1.14 | 3-1 | 7.60 | 2.79 | 6-11 | 7.20 | 2.17 |
| Good News Network | 5 | 21 - 31 | 25.60 | 4.16 | 6-11 | 8.60 | 1.82 | 5-11 | 7.60 | 2.41 | 8-11 | 9.40 | 1.14 |
| **Evolve** | 3 | 24 - 27 | 25.00 | 1.73 | 9-11 | 9.67 | 1.15 | 4-9 | 7.33 | 2.89 | 6-9 | 8.00 | 1.73 |
| **DBT Self-Help** | 2 | 25 - 29 | 27.00 | 2.83 | 8-11 | 9.50 | 2.12 | 9-11 | 10.0 | 1.41 | 6-9 | 7.50 | 2.12 |
| **Action Mental Health website** | 2 | 23 - 27 | 25.00 | 2.83 | 9 | 9.00 | 0.00 | 7-9 | 8.00 | 1.41 | 7-9 | 8.00 | 1.41 |
| **HSE Mental Health website** | 1 | 21 |  |  |  |  |  |  |  |  |  |  |  |
| **Insight Timer** | 1 | 27 |  |  |  |  |  |  |  |  |  |  |  |
| **Spunout Empathy Online (unit within spunout Academy)** | 1 | 23 |  |  |  |  |  |  |  |  |  |  |  |
| **Student Health App** | 1 | 23 |  |  |  |  |  |  |  |  |  |  |  |
| **Overall** | **26** | **18 - 31** | **24.73** | **3.02** | **4-11** | **8.76** | **1.50** | **3-11** | **7.96** | **1.89** | **6-11** | **8.02** | **1.55** |

Behavioural engagement was high across the tools and among the participants (M = 8.76, SD = 1.5). Whereas cognitive engagement (Mean = 7.96, SD = 1.89), indicated more mental effort and larger variance among participants. Intrusive ads, repetitive logging, long interactions, and paywalls disrupted engagement and contributed to attrition. These barriers were described as frustrating, demotivating, or “breaking the flow,” underscoring the importance of low-friction design. Tools that demanded sustained attention or repeated effort were quickly abandoned, even when participants recognised their potential value.

### CFIR framework

The findings are organised using the four CFIR domains most relevant to digital mental health implementation: Intervention Characteristics, Outer Setting, Characteristics of Individuals, and Process. Quantitative SUS (Table 5) and TWEETS (Table 6) scores are woven throughout to illustrate how implementation determinants manifested in measurable usability and engagement outcomes. The CFIR framework provided a structured lens for interpreting how usability, trust, cultural relevance, emotional needs, and behavioural reinforcement shaped implementation potential.

#### Intervention characteristics

Usability was the most influential intervention characteristic shaping acceptability (Table 5). Just two tools, spunout Navigator and the Student Health App, exceeded the SUS benchmark, and these were consistently described as simple, clear, and easy to navigate. Participants emphasised that tools requiring minimal steps, offering clean layouts, and avoiding cognitive overload were the ones they were most likely to use. In contrast, tools with lower SUS scores were described as overwhelming, too long, or requiring “too many clicks,” which discouraged continued use.

Adaptability and personalisation also shaped perceptions of intervention quality. Tools such as Bearable and Insight Timer were valued for allowing users to tailor content, track mood, or adjust complexity. Participants described gaining insight into emotional patterns and appreciated tools that reflected their identities or contexts. However, excessive options created cognitive overload, reinforcing the need for manageable rather than maximal personalisation.

Perceived evidence, strength, and credibility further influenced acceptability. Tools linked to the HSE, NHS, Action Mental Health, or universities were interpreted as trustworthy and safe. Participants described being more inclined to use tools “backed up by research” or associated with recognised institutions. Commercial apps generated cautious trust, with concerns about profit motives and data practices. However, credibility checks were not universal, particularly for positive-content tools where emotional benefit outweighed evidence considerations.

#### Outer setting

Institutional trust and governance clarity were central to how young people interpreted the legitimacy of digital mental health tools. National health organisations, universities, and established charities provided a sense of safety and reliability, while commercial platforms were approached with scepticism. This reliance on institutional markers reflects the importance of external validation in shaping adoption.

Peer influence also played a significant role. Participants valued hearing from “people my age,” and youth-led design or testimonials increased trust and relatability. Peer perspectives acted as informal evidence, complementing formal institutional endorsement. This dual trust pathway, formal and informal, was particularly important for marginalised youth who often rely on social networks to navigate digital spaces.

Cultural relevance further shaped acceptability. Participants highlighted the need for content that reflected their lived experiences, identities, and local contexts. Tools lacking representation or culturally grounded examples were perceived as less relatable, reducing their perceived usefulness.

#### Characteristics of individuals

Young people’s engagement with digital tools was shaped by their emotional needs, expectations, and digital health literacy. Participants demonstrated sophisticated critical literacy, routinely checking creators, evidence bases, privacy policies, and update frequency. However, emotional needs sometimes overrode critical appraisal. Positive-content tools were used primarily for mood lifting, distraction, and grounding.

Young people did not approach digital tools as clinical interventions. Instead, they used them as light-touch emotional coping supports. This distinction helps explain why tools with lower usability still achieved high engagement. Tools that felt “school-like” or “clinical” were rejected, even when they were evidence-based.

#### Process

Engagement and habit formation were shaped by behavioural cues such as reminders, streaks, short tasks, and predictable interfaces. These mechanisms aligned closely with TWEETS engagement patterns (Table 6). Participants described notifications as helpful for maintaining routines, while variety and novelty supported sustained attention. Conversely, long interactions, repetitive logging, intrusive ads, and paywalls disrupted engagement and contributed to attrition.

Execution was influenced by the degree to which tools supported low-effort, everyday use. Tools that opened immediately, required no sign-up, and used simple language were more likely to be integrated into daily routines. In contrast, tools requiring repeated log-ins, complex navigation, or extended tasks were quickly abandoned.

Across CFIR domains, several constructs emerged as particularly salient. Within Intervention Characteristics, usability, simplicity, and adaptability were central to acceptability. In the Outer Setting, institutional trust, perceived safety, and concerns about data governance shaped legitimacy. Characteristics of Individuals highlighted emotional readiness, digital literacy, and the need for culturally grounded content. Within Process, habit formation, behavioural reinforcement, and clear signposting influenced sustained engagement. These constructs interacted to create a coherent implementation pathway in which usability and trust functioned as foundational determinants, while cultural relevance and emotional needs shaped depth and continuity of engagement.

## Discussion

This study provides one of the first all-island examinations of the acceptability and implementation potential of digital mental health tools among marginalised young people in Ireland, integrating quantitative usability and engagement data with in-depth qualitative insights. The CFIR-aligned findings demonstrate that implementation is shaped by the interaction of intervention characteristics, contextual influences, individual needs, and engagement processes.

Usability emerged as the strongest determinant of acceptability in shaping young people’s ongoing interaction with tools, but credibility emerged as the strongest determinant of *initial adoption*. Just two tools, spunout Navigator (M = 71.88) and the Student Health App (M = 69.58), exceeded the SUS benchmark of 68, and participants consistently described these tools as simple, clear, and easy to navigate. Tools with lower SUS scores were described as overwhelming, cluttered, or requiring too many steps, which discouraged continued use. These qualitative accounts closely mirrored the quantitative usability patterns, reinforcing that usability is a critical gateway to engagement, consistent with previous research exploring usability of a prototype digital mental health platform with students [26]. This also aligns with Schueller et al. [27], who found that preferences for simplicity versus detailed tracking vary widely, indicating that usability is not only about ease but also about cognitive fit.

However, usability alone did not determine engagement. The TWEETS results showed that tools such as Bearable (M = 25.88) and Good News Network (M = 25.57) achieved high engagement despite lower usability. Qualitative findings explain this divergence: young people used digital tools primarily for emotional regulation, distraction, grounding, and/or mood lifting rather than structured therapeutic intervention. Tools that provided immediate emotional payoff were used more consistently, even when their usability was imperfect. This distinction between usability and emotional resonance reflects broader evidence that engagement is shaped by affective reward rather than design quality alone [28,29]. Ho et al. [30] similarly found that young people often prioritise emotional benefit over interface quality when selecting tools for everyday coping.

Credibility and trust were central to initial adoption. Participants relied heavily on institutional affiliation, particularly with the HSE, Action Mental Health, NHS, or universities, to determine whether a tool was safe and evidence based. This demonstrates that *credibility*, defined as perceived legitimacy, safety, and trustworthiness, is the primary filter through which young people decide whether a tool is worth trying at all [8]. Peer perspectives also acted as informal evidence, with young people valuing reviews, testimonials, and youth-led design. These findings strongly echo Ho et al. [30], who reported that young people were more likely to use a tool if it was funded by a reputable organisation, advertised on a verified page, or recommended by trusted peers. At the same time, credibility checks were not universal: for positive-content tools, emotional benefit outweighed evidence, and participants openly acknowledged not checking sources when the content simply made them feel better. Topooco et al. [31] and Madrid-Cagigal et al. [32] similarly found that students were more familiar with general wellbeing apps than clinically oriented Digital Mental Health Interventions (DMHIs), suggesting that credibility matters most when tools are perceived as “treatment” rather than “support.” Taken together, these findings clarify the distinction between *credibility* and *usability*: credibility determines whether young people will *start* using a tool, while usability determines whether they will *continue* using it. Credibility acts as a gatekeeper for adoption; usability acts as a gatekeeper for sustained engagement.

Personalisation and self-monitoring enhanced relevance and supported self-insight. Tools that allowed users to tailor content, track mood, or adjust complexity were perceived as more useful. This aligns with Schueller et al. [27], who noted wide variation in preferences for personalisation and simplicity, and with Riboldi et al. [33], who reported that lower personalisation compared to in-person support was a key disadvantage for students. However, excessive options created cognitive overload, particularly for neurodivergent participants, highlighting the need for manageable rather than extensive personalisation. Cross et al. [34] similarly emphasise that personalisation must be “scaffolded” rather than limitless to avoid overwhelming young users. Cultural and contextual relevance also shaped acceptability, with participants expressing a desire for more Irish or UK-specific content and examples that reflected their lived experiences.

Engagement processes were shaped by behavioural cues such as reminders, streaks, short tasks, and predictable interfaces. These mechanisms aligned closely with TWEETS engagement patterns and helped young people integrate digital tools into their daily routines. This is consistent with Saleem et al. [29], who identified tailored reminders as an effective engagement strategy, and with Jackson et al. [35], who found that students preferred short, predictable, low-effort digital interactions. Conversely, long interactions, repetitive logging, intrusive ads, and paywalls disrupted engagement and contributed to attrition. These findings demonstrate that sustained engagement depends on a combination of emotional payoff, usability, and behavioural reinforcement.

Overall, the findings show that digital mental health tools are experienced not only as technological products but as interventions embedded within broader social and emotional contexts. Their acceptability and implementation potential depend on credibility, usability, trust, cultural relevance, emotional resonance, and the extent to which they support low-effort, everyday use. This aligns with Lipschitz et al. [28], who argue that engagement challenges reflect deeper motivational and contextual factors rather than design issues alone.

These findings have several practical implications. Developers need to prioritise low-friction design, clear navigation, and culturally grounded content that reflects the identities and lived experiences of diverse youth. Institutional affiliation and transparent data practices are essential for building trust. Youth workers and community organisations can play a key role in supporting digital literacy, facilitating safe engagement, and integrating digital tools into everyday youth-centred settings. Service providers should avoid overwhelming young people with multiple tools and instead focus on simple, coordinated pathways that reduce fragmentation.

### Strengths

This study has several strengths. It provides one of the first all-island analyses of digital mental health implementation among marginalised youth in Ireland, integrating quantitative usability and engagement metrics with qualitative thematic analysis. The mixed-methods design enabled triangulation across behavioural, experiential, and implementation-focused data. Co-design with the Youth Reference Group enhanced cultural relevance and ecological validity. The inclusion of diverse youth — including migrants, asylum seekers, LGBTQ+ young people, and neurodivergent participants — strengthens the study’s contribution to equity-focused digital mental health research.

### Limitations

This study has several limitations that should be considered when interpreting the findings. The sample size, while appropriate for mixed-methods research, limits generalisability as previously established [36]. The final survey included 24 participants, and engagement data were tool-specific, with some tools having very small user numbers (e.g., n = 1–2). This uneven distribution may have influenced mean SUS and TWEETS scores, particularly for less frequently used tools.

The two-week trial period may not fully capture long-term engagement or attrition patterns typical of digital mental health use. Digital tools often experience steep drop-off after initial use, and a longer study period may have revealed different engagement trajectories, consistent with [12,37]. Self-selection bias may also have influenced participation, as young people with higher digital literacy or interest in mental health may have been more likely to take part [12].

Although recruitment prioritised marginalised youth, representation varied across groups, and some communities, such as Traveller youth and newly arrived migrants, may remain underrepresented. The presence of responses from young people not resident in Ireland or otherwise not meeting the selection criteria for this study in the pre-workshop dataset required data cleaning and may have influenced early patterns of tool use. Finally, the study evaluated tools in a real-world, uncontrolled environment, meaning usage was not standardised. While this enhances ecological validity, it limits the ability to compare tools under consistent conditions.

The findings highlight several gaps that future research should address to strengthen the evidence base for youth-centred digital mental health implementation. First, longitudinal studies are needed to examine long-term engagement trajectories, including how patterns of use evolve beyond short trial periods and which design features support sustained interaction over months rather than weeks. This is particularly important given the study’s finding that emotional resonance can override usability in the short term, suggesting that different mechanisms may drive early versus sustained engagement.

Further, future work should investigate how digital mental health tools can be embedded sustainably within youth-centred settings, such as youth work organisations, community groups, and educational environments. Implementation studies in these settings could clarify how digital supports complement existing relational, social, and practical supports, and how youth workers can act as trusted intermediaries who enhance credibility and uptake.

In addition, targeted research with specific marginalised groups—including migrant youth, Traveller communities, LGBTQ+ young people, neurodivergent youth, and young people with disabilities—is essential to understand culturally specific needs, preferences, and barriers. The present study indicates that cultural relevance and institutional trust strongly shape acceptability; future research should therefore explore how co-design, representation, and culturally grounded content influence credibility and engagement across diverse youth populations.

Further investigation is also needed into moderated peer-support models, which may offer a credible and relatable alternative to purely clinical or commercial tools. Research should examine how peer moderation affects trust, safety, and emotional resonance, and whether hybrid models combining peer support with professional oversight can enhance acceptability among marginalised youth.

Future studies moreover should evaluate digital literacy interventions that aim to strengthen young people’s ability to assess credibility, navigate digital tools, and make informed choices. Although digital literacy was high in this sample, it remains uneven across youth populations, and targeted interventions may help reduce inequities in digital engagement.

Finally, research should focus on governance frameworks that enhance trust, transparency, and safety. Young people in this study relied heavily on institutional markers of legitimacy; therefore, future work should examine how accreditation systems, data protection standards, and youth-friendly governance models influence adoption and sustained use.

Together, these research directions can support the development of digital mental health ecosystems that are trustworthy, culturally relevant, and responsive to the needs of diverse youth communities.

### Recommendations

The findings of this study indicate that the designers and commissioners of digital mental health tools for marginalised young people must prioritise usability, credibility, cultural relevance, and emotional resonance to support adoption and sustained engagement. Only two tools surpassed the SUS usability benchmark, albeit based on a small user sample, indicating that future development and commissioning should emphasise simple, mobile-first, low-friction design without long pages, excessive options, repeated logging, or complex navigation. Usability should be treated as a core requirement rather than an optional enhancement.

Credibility must be communicated clearly. Participants relied heavily on institutional affiliation, such as the HSE, NHS, Action Mental Health, or universities, to determine whether a tool was trustworthy. Developers and service providers should therefore make evidence bases, affiliations, and governance structures visible and accessible. At the same time, peer perspectives were powerful motivators, suggesting that youth-generated reviews, testimonials, and co-designed content may enhance trust and relatability.

Cultural and contextual relevance should be strengthened. Participants expressed a desire for content that reflected their identities, lived experiences, and local contexts, including Irish and UK examples. Tools that lacked representation or cultural grounding were perceived as less relatable. Codesign with diverse youth communities, including migrants, LGBTQ+ youth, and neurodivergent young people, may help ensure that digital tools reflect the realities of those they aim to support.

Emotional relevance should be recognised as a legitimate mechanism of engagement. Young people used digital tools primarily for mood lifting, distraction, and grounding rather than structured therapeutic intervention. Tools that provide immediate emotional payoff may, therefore, be more acceptable and more likely to be used consistently. It is important for developers to consider how design, tone, and content can support emotional regulation without overwhelming users.

Finally, engagement features such as reminders, streaks, and short tasks should be used judiciously to support routine use without creating pressure or fatigue. Intrusive ads, paywalls, and long interactions were consistently described as barriers and should be avoided wherever possible.

## Conclusion

In conclusion, digital mental health tools hold meaningful potential for supporting marginalised young people when they are simple, trustworthy, culturally relevant, and aligned with everyday contexts. Usability and trust act as foundational determinants of engagement, while emotional resonance and behavioural reinforcement shape sustained use. Implementation strategies must prioritise clarity, safety, and youth-centred design to ensure equitable access and meaningful impact.

Taken together, these findings indicate that effective implementation will require coordinated cross-sector collaboration across statutory services, youth work, community organisations, migrant and cultural groups, and lived-experience networks. No single sector can deliver the breadth of supports required to meet the diverse needs of young people.

## Data Availability

The data underlying this study cannot be shared publicly due to ethical restrictions. The participant group is small and includes marginalised youth, and sharing individual?level data would pose a risk of re?identification and compromise participant safety. All relevant materials needed to understand and evaluate the study (including analytic procedures and supporting information) are provided within the manuscript and its supplementary files. Additional non?identifiable information may be made available upon reasonable request to the corresponding author, subject to institutional ethical approval.

## Acknowledgements

The authors would like to acknowledge the funders who supported this work as well as Jamie McNulty as part of this research project and the contribution of the Reference Group, the young people who took part in the study, and the organisations that facilitated their participation. The authors would also like to thank Joanne Kearney and Conor Ross of GlowMetrics for the setup and configuration of Google Data Studio to capture the study usage data.

## Supporting Information Captions

S1 Appendix. **Details on individual digital tools included in evaluation**

S2 Appendix. **Question prompts for workshops/individual interviews**

S3 Appendix. **Acceptability and implementation questions in final survey**

## Notes

### Competing Interest Statement

The authors have declared no competing interest.

### Author Declarations

The Ethics Committee of Ulster University in Northern Ireland gace full ethical approval for this work (FCPSY-25-027-A)

